# Comparison of ultrasound-guided erector spinal muscle plane block and quadratus block for laparoscopic renal cancer resection:A single-center,double-blind, randomized controlled trial

**DOI:** 10.1101/2024.03.01.24303596

**Authors:** Meng Zhang, Shuchuan Zhao, Mingfang Li, Yue Liu, Hu Li, Peng Su, Guangmin Xu

## Abstract

**Objective:** This study investigated the effects of ultrasound-guided erector spinal muscle plane block (ESPB) and quadratus muscle block (QLB) on the quality of analgesia and recovery after laparoscopic nephrectomy.

**Design:** randomized, controlled, double-blind study.

**Setting:** A single tertiary care academic medical center,include anesthesia preparation room, operating room, anesthesia recovery room and ward.

**Patients:** Aged 18-70years,ASA grades I-III,elective laparoscopic partial nephrectomy or radical nephrectomy and 54 patients were included in the statistical analysis.

**Interventions:** All included patients were randomassigned to the erector spinal muscle plane block or the quadratus block,and all patients underwent morphine pump controlled analgesia.

**Results:** The study found that ultrasound-guided ESPB had a higher incidence of hypotension than QLB at the T1 time point, but it did not significantly increase the intraoperative dose of the vasoactive drug used. Patients in the ESPB group showed significant improvement in resting NRS pain scores at 0.5h,number of morphine pumps at 6h and 24h, cumulative morphine equivalent consumed 6h after surgery, and QOR-15 score at 24 h after surgery, and shortened hospital stay.

**Conclusions:** Compared with QLB,ESPB has certain advantages in analgesia and recovery quality after laparoscopic nephrectomy, and shows opioid frugality effect at individual postoperative time points.

**Trial registration:** The trial was registered prior to patient enrollment at the Chinese Clinical Trial Registry on 15/08/2023 (ChiCTR2300074743).

**Key point:**

1. Although regional anaesthsia is beneficial for laparoscopic renal cancer resection, it is still controversial which type of regional anaesthsia is most appropriate.
2. A good regional anaesthsia not only has a long postoperative analgesia time, but also has better anesthesia recovery quality.
3. The primary outcome measure was the cumulative consumption of morphine equivalent within 6h after surgery.
4. ESPB is a good regional anaesthsia that can be used for laparoscopic renal cancer resection.
5. In this randomised controlled trial of patients undergoing laparoscopic renal cancer resection, compared with QLB, ESPB has certain advantages in analgesia and recovery quality after laparoscopic nephrectomy.

## 1. Introduction

Malignant kidney tumors account for 2-3% of the global cancer burden and their incidence is increasing[1].It is reported that more than 430,000 people were diagnosed with kidney cancer worldwide in 2020 and led in more than 170,000 deaths [2], and surgery is currently the most important treatment for kidney cancer[3]. The incidence and degree of acute pain in the early stage after laparoscopic renal cancer resection are not significantly different from that of open surgery. Moderate to severe pain in the early postoperative period [4]. Poor pain control will lead to the occurrence of neuroendocrine stress response, cardiovascular and cerebrovascular events, decreased immune function, and increased risk of infection related [5][6].

At present,the main analgesia of postoperative pain [7] are oral NSAIDs drugs,intravenous opioid analgesia,and regional nerve block analgesia. For example,oral NSAIDs drugs may increase the risk of gastrointestinal bleeding[8],and the side effect of opioids(e.g.nausea,vomiting,itching,respiratory depression,intestinal obstruction)limit the dosage of drugs[9], affecting the analgesic effect and functional recovery. Regional nerve block analgesia(e.g.epidural block, thoracic paravertebral block) as part of the multimodal analgesia technique in postoperative pain management of renal cancer has insufficient[10,11], hematoma, pneumothorax, hypotension and so on.

Ultrasound-guided quadratus muscle block (QLB) and erector spinal muscle plane block (ESPB) are simple and novel fascia plane block for postoperative analgesia in patients undergoing laparoscopic nephrectomy (LN)[12,13], but there is no randomized controlled study comparing the difference between the two methods in LN surgery. This study compared the two methods to provide the basis for LN anesthesia and postoperative analgesia.

## 2. Methods

This single-center, randomized, controlled,double-blind study was prospectively registered at the Chinese Clinical Trial Registry(ChiCTR2300074743) through the Sichuan Provincial People’s Hospital.Our study methods were performed in accordance with the guidelines and regulations of the clinical registry.

Patients undergoing elective laparoscopic partial, total or radical renal cancer resection from August 2023 to October 2023 were selected, aged 18 to 70 years, body mass index (Body Mass Index, BMI) 18 to 30 kg/m2, American Association of Anesthesiologists (American Society of Anesthesiologists, ASA) grade I-III. Exclusion criteria for preoperative elevated intracranial pressure, motion sickness, glaucoma, severe hypertension or other diseases causing nausea and vomiting, unable to cooperate with communication and operation, platelet or coagulation function abnormal, local anesthetic allergy history, puncture area infection, serious heart, lung, brain, liver and kidney function abnormalities, intraoperative changes, chronic opioid addiction or use other analgesic drugs for more than 3 months.

Using SPSS 25.0 (IBM, Chicago,IL,USA) software produces a random number list, Patients 1:1 into two groups, Group ESPB and QLB, An investigator blinded to the study content and grouping prepared an opaque envelope for each patient, Patients, 1h before surgery, The anesthesiologist performed ultrasound-guided erector spinal muscle plane block (erector spinae plane block, ESPB) or squastus muscle block (Quadratus Lumborum Block, QLB), Anesthesia was performed by another anesthesiologist, The surgical team, nursing team, data collectors, and statistical analysts were blinded to the grouping.

### 2.1. Ethics

Ethical approval for this study was provided by the Ethics Committee of Sichuan Academy of Medical Sciences, Chengdu, Sichuan (Chairperson Prof Liangping Li)on 19 January 2023,Approval Number: LunShen No.24,2023.Patients and family members signed informed consent before enrollment.The CONSORT flow diagram was used for enrollment and allocation of patients (Fig.1).

**Fig.1.**
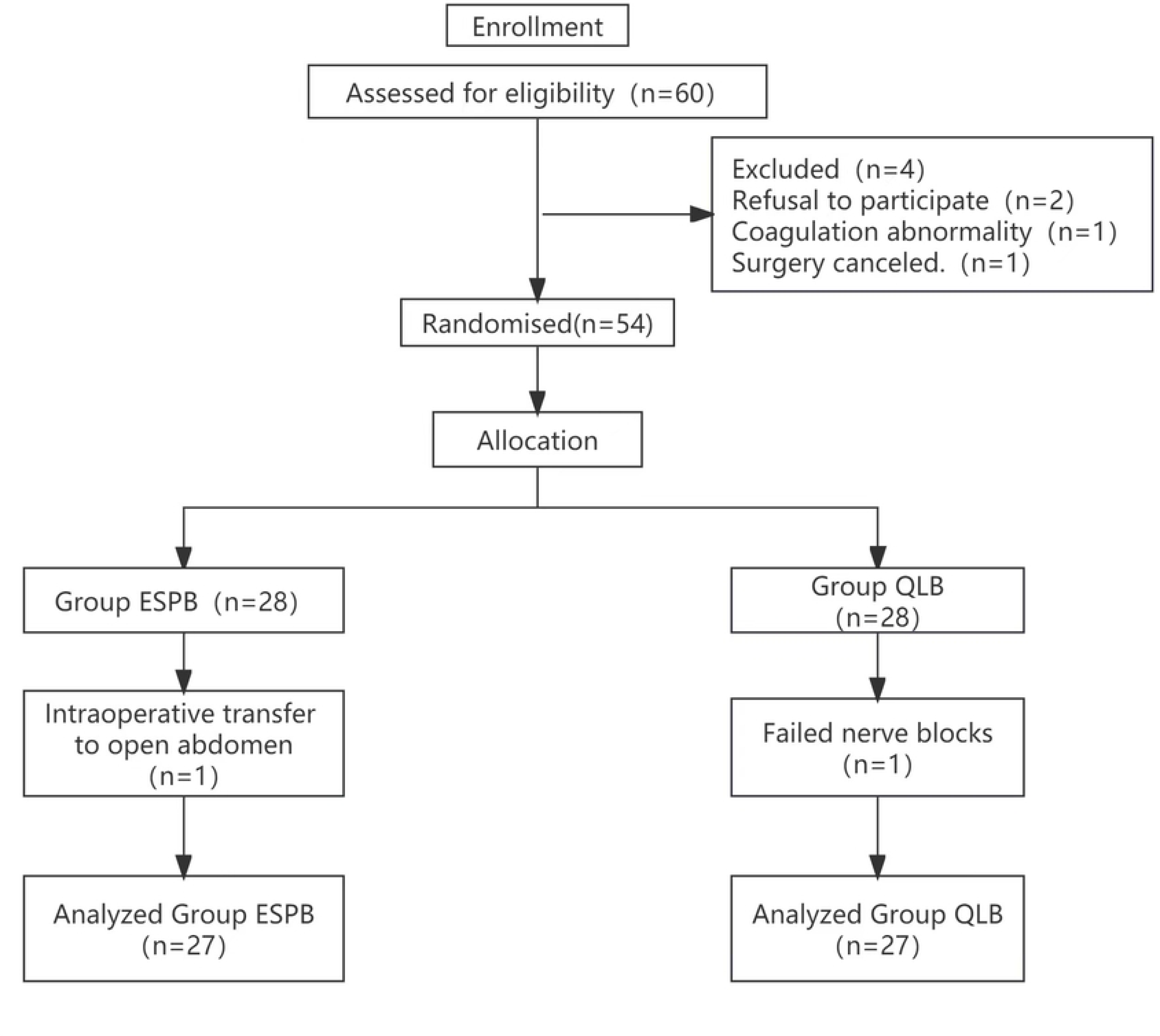
Consort flow diagram of patients.

### 2.2. Surgical technique

The laparoscopic surgery was performed by the members of the same surgical team. For the retroperitoneal approach procedure, the patient was placed in the lateral decubitus position, requiring three puncture incisions. The first lies below the 12th rib of the posterior axillary line; the second lies 2cm above the iliac crest of the central axillary line, and the third lies below the costal margin of the axillary front. In the case of radical nephrectomy, the first incision extends ventrally to remove the kidney, with the remaining two incisions positioned unchanged. For surgery with the transperitoneal approach, the patient was placed in a semi-oblique decubitus position with three or four trocars distributed between the navel and xiphoid process from the abdominal midline to the axillary front. The pneumoperitoneum pressure was maintained at 12-16 mmHg throughout the surgical procedure.

### 2.3. Block procedures

Open peripheral vein of upper limbs, monitoring HR, BP, ECG, SpO 2, invasive radial artery puncture and monitoring. All nerve blocks were completed by the same anesthesiologist with 3 years of experience in nerve block, high-frequency line array or low-frequency convex array probe (2-13 MHz, M-Turbo, FUJIFILM SonoSite, Inc, USA) and 22-gauge nerve block needle (80 mm B.Braun Meisungen AG, Germany) were used.

#### 2.3.1. US-guided ESPB

The patient used the lateral position of the patient, moving the probe from the 12 ribs to the tip of the T 10 transverse process, using the out-of-plane technology into the needle, the needle reached between the vertical spinal surface of the transverse process surface, injection of 0.9% normal saline 1∼2ml to confirm the tip position, 0.4% ropivacaine 25ml, the transverse process and the erector spinal muscle were separated by local anesthetic.(Fig.2.)

**Fig.2.**
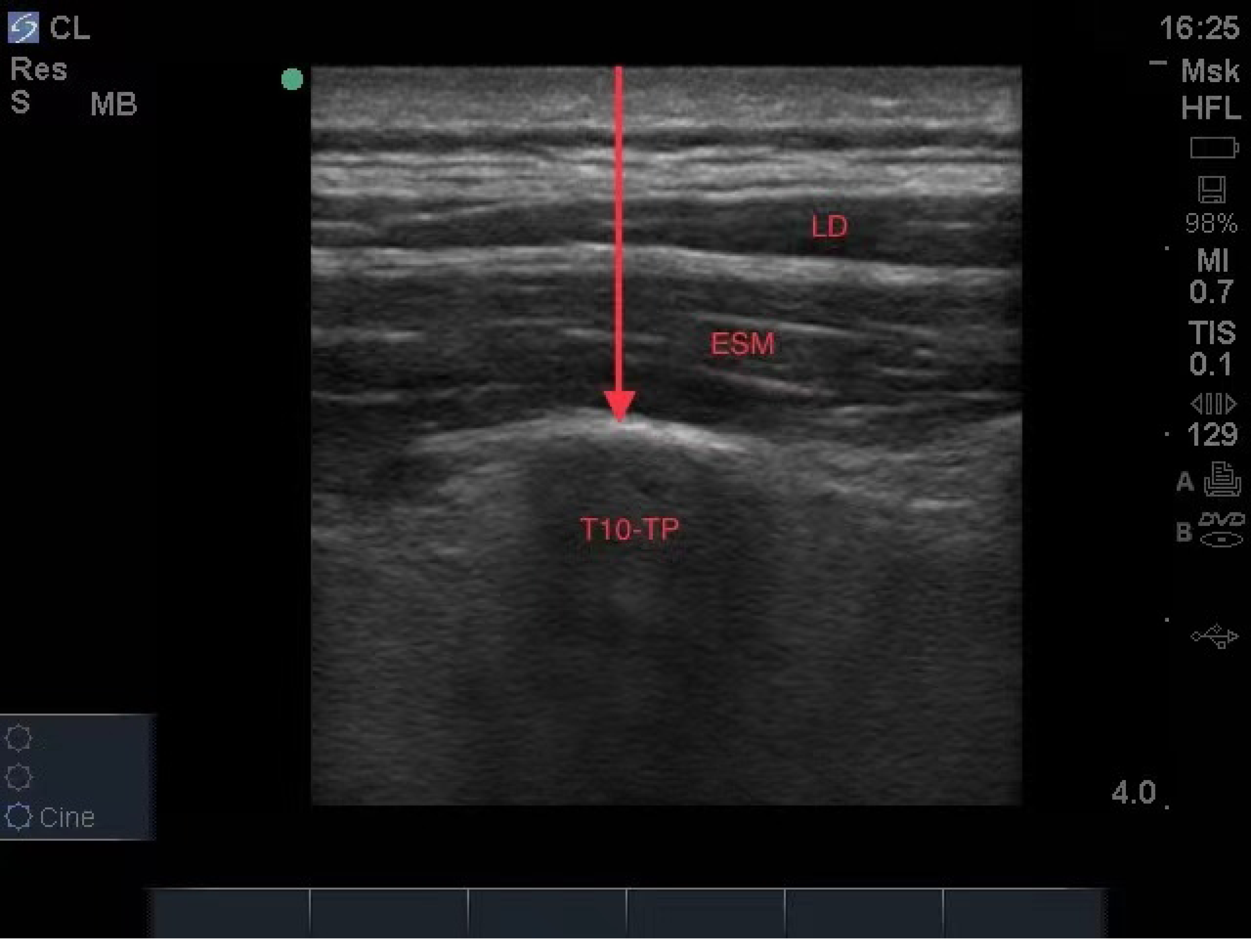
Relevant sonoanatomy for US-guided ESPB. Relevant sonoanatomy for US-guided ESPB. ESM = erector spinae muscle; LD = latissimus dorsi;TP=transverse process.

#### 2.3.2. US-guided QLB

The patient was in the lateral position, lateral up, disinfected and spread, and using a low frequency line array ultrasound probe, the probe was placed axially on the L3 level and the iliac arch and moved backward until the quadratus muscle and psoas muscle on the superficial surface of the transverse us abdominis were seen.After real-time ultrasound-guided in-plane injection, a needle was inserted from the dorsal side to the ventral side, and injection of 0.9% saline, 25 ml was injected into the posterior side of the quadratus muscle, the medial edge of the erector spinal muscle, the inferior edge of the latissimus dorsal muscle, and the triangle of the lumbar fascia.(Fig.3.)

**Fig.3.**
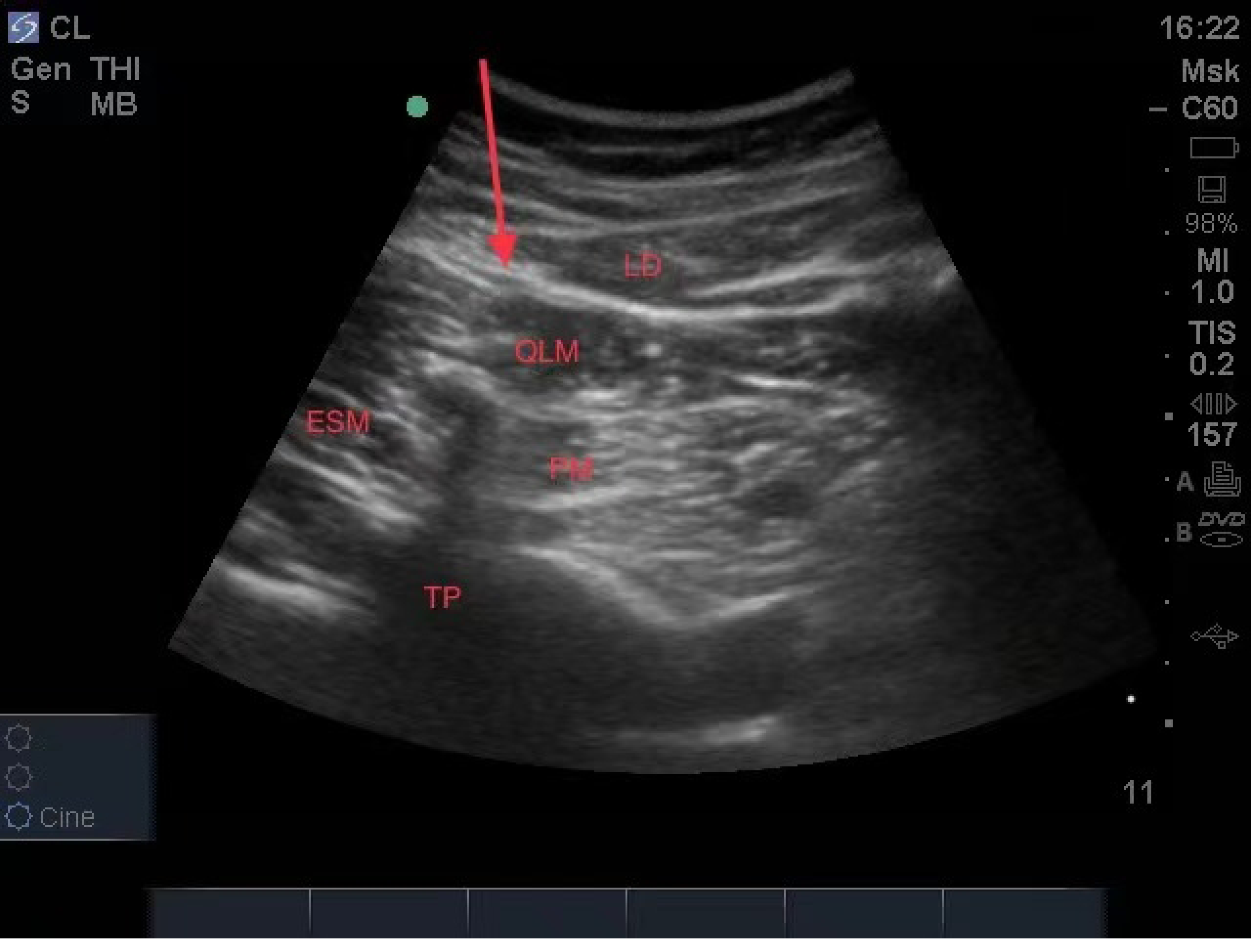
Relevant sonoanatomy for US-guided QLB. Relevant sonoanatomy for US-guided QLB. LD = latissimus dorsi; QLM = quadratus lumborum muscle; ESM = erector spinae muscle; PM = psoas muscle;TP = transverse process.

#### 2.3.3. Determination of the range of the nerve block

Measurement of the range of nerve block are by an anesthesiologist blinded to the experimental method, 30min after the completion of the block, using the alcohol contact skin cold extinction method (with alcohol cotton ball contact block area skin, cold decreased even disappear, rather than the block area can feel cool alcohol) and record the block range, 30 min in the operation area skin cold decreased as block failure.

### 2.4. Perioperative analgesia and management

#### 2.4.1. Anesthesia management

After admission to the operation room, all patients received standard monitoring, including ECG,blood oxygen saturation,invasive artery monitoring,electroEEG monitoring,body temperature monitoring,general anesthesia with endotracheal intubation,midazolam 0.04mg/kg,sufentanyl 0.3μg/kg,cis atracurium 0.15mg/kg, propofol 2mg/kg.Anesthesia maintenance:remifentanil 0.1-0.2μg/kg·min, propofol 4-12 mg/kg·h for CSI 40 to 60, infusion speed of remifentanil l and propofol was adjusted for mean arterial pressure and heart rate (within±20% of preoperative values).10 to 15min before the end of the surgical intravenous injection of sufentanil 0.05 ug/kg to prevent postoperative pain, 5mg of intravenous injection to prevent postoperative nausea and vomiting, all anesthetic drugs were discontinued at the end of the surgical suture, and the controlled intravenous analgesia pump: (morphine hydrochloride 50mg+tropisetron 5mg+0.9% sodium chloride injection diluted to 100ml, single press dose of 4ml, locking time 15min). After the patient is removed,the anesthesia recovery room for further monitoring. If the NRS pain score is 4, 5 ug of sufentanil will be given IV,and repeated if necessary,the patient will be transferred to the anesthesia recovery room.

#### 2.4.2. Intraoperative treatment

When the MAP decreased less than 20% of the basal value,the intravenous ephedrine 6mg or noradrenaline 50 ug, and the increase of MAP was greater than 20% of the basal value,the intravenous rapid infusion of remifentanil 40 ug+propofol 30mg showed no obvious effect after 1min,and intravenous nocardipine 0.1mg. IV atropine 0.3 mg for HR less than 50 times / minute, and intravenous esmolol 10 mg for HR greater than 100 times / minute. All of the above drugs can be repeated if necessary.

#### 2.4.3. Analgesia management

Patients were selected according to the inclusion exclusion criteria,Detailed explanation of the pain scores to the patient Numeric Rating Scale(NRS) definition and the Patient-Controlled Intravenous Analgesia(PCIA) for the method of use, The NRS pain score represents the degree of pain by 0 to 10, (0 To a painless, 10 represents the most painful, Patients pick a number to represent their level of pain), NRS pain score 4, Patient press-controlled analgesia pump, Patients based on the pain relief, Repeated pressing (automatic analgesia pump locking time of 15min, No additional morphine consumption is added during this period), No relief of the pain after 5 min, Intravenous tramadol 100mg or desoxin 5mg for analgesia.

### 2.5. Outcome measurements

The primary outcome measure was the cumulative consumption of morphine equivalent within 6h after surgery.

Secondary outcome measures are the duration of nerve block operation (between the tip of nerve block needle to the end of ropivacaine local anesthetic), the depth of nerve block puncture (the distance between the tip of nerve block needle and the injection of local anesthetic). MAP and HR during the anesthesia preparation (T0), 5min (T1), 5min (T2), 5min before tracheal extubation (T3), 5min (T4), and leaving the PACU (T5). The amount of intraoperative vasoactive drugs used. Intraoperative remifentanil and propofol dosage; operation time and anesthesia time. NRS pain scores at rest and cough at 0.5h / 6h / 24h / 48h after surgery; scale (QoR-15 at 24h / 48h), nerve block-related complications (hematoma at the puncture site, puncture site infection, local anesthetic poisoning, respiratory depression, nausea and vomiting, lower limb weakness on the affected side, etc.); first exhaust time; first time of ambulation; postoperative hospital stay.

### 2.6. Sample size calculation

According to the preliminary pilot study of 10 patients in each group (QLB, 6.63 ±4.61mg;ESPB,3.51±3.25mg), both groups consumed the cumulative morphine equivalent within 6 hours after surgery to determine the sample size. Sample size was calculated using two independent sample t-tests and using the software (G*power v3.1.9.7,Germany), α is 0.05, control (test efficacy) 1- β is 80%, and effect size is 0.78. At least 54 cases with 27 observed values in each group were required for this study. Considering the 10% loss-up rate and refusal, 60 patients were included in this study.

### 2.7. Statistical analysis

In this study, the statistical analysis was performed in the SPSS22.0 software, Quantitative data of a normal distribution,The data are described using the mean and the standard deviation, Quantitative data of the non-normal distribution, Data are described using the median and interquartile spacing;Categorical variables are expressed in the rate or composition ratio; The quality of variance of quantitative data with normal distribution was determined by t test, If the variance is uneven, The t’test was used; Quantitative data were not normally distributed using the non-parametric wilcoxon rank-sum test; A Chi-square test was performed for the categorical variables, When the theoretical frequency of over 20% of cells is less than 5, Using the Fisher exact probability method;All statistical tests were two-sided,P<0.05 will be considered statistically significant.

## Results

A total of 60 patients were included in this study,including 2 refused to participate, 2 cancelled, 1 transferred open, and 1 nerve block failure. Finally,54 patients were included in the statistical analysis, 27 patients in each group in Fig 1.

There were no statistical differences in gender, age, height, BMI, ASA grade, hypertension, surgical history, surgical location, surgical approach, surgical resection range, surgical duration,amount of propofol and remifentanil,anesthesia duration and preoperative QoR-15 score (p>0.05)(Table 1).

**Table 1.** Demographic and perioperative characteristics.

|  | QLB (n = 27) | ESPB (n = 27) | p value |
| --- | --- | --- | --- |
| Age (year) | 48.48 ± 11.26 | 50.04 ± 8.33 | 0.822<sup>#</sup> |
| Sex |  |  | 0.584<sup>&</sup> |
| M | 16 | 14 |  |
| F | 11 | 13 |  |
| Height (cm) | 163.56 ± 9.12 | 162.11 ± 7.817 | 0.535<sup>*</sup> |
| BMI (kg/m<sup>2</sup>) | 22.95 ± 2.43 | 24.30 ± 2.99 | 0.074<sup>*</sup> |
| ASA |  |  | 0.340<sup>&</sup> |
| I | 5 | 8 |  |
| II | 22 | 19 |  |
| Hypertension | 4 (14.8%) | 5(18.5%) | 1<sup>&</sup> |
| History of surgery | 14 (51.9%) | 12 (44.4%) | 0.586<sup>&</sup> |
| Surgical location |  |  |  |
| Left | 16 | 15 | 0.783<sup>&</sup> |
| Right | 11 | 12 |  |
| Operative approach |  |  | 0.34<sup>&</sup> |
| Belly | 19 | 22 |  |
| Retroperitoneum | 8 | 5 |  |
| Excision extension |  |  | 0.738<sup>&</sup> |
| Ablatio | 17 | 16 |  |
| All excision | 7 | 6 |  |
| Radical correction | 3 | 5 |  |
| Preoperative QoR-15 score | 146.7±4.82 | 147.56±4.76 | 0.327<sup>#</sup> |
| The duration of the procedure (min) | 100.48±46.44 | 96.15±32.28 | 0.755<sup>#</sup> |
| Duration of anesthesia (min) | 119.15±48.11 | 116.01±35.06 | 0.802<sup>#</sup> |
| Amount of propofol (mg) | 643.19±313.99 | 589.18±251.35 | 0.829<sup>#</sup> |
| The dosage of remifentanil | 813.97±337.63 | 759.06±337.35 | 0.672<sup>#</sup> |

Compared with patients in the QLB group, the EPSB group decreased within 6h, 24h and 48h after surgery, with a statistical difference in morphine equivalent consumption within 6h (P <0.05) and no statistical difference at other time points (P> 0.05) (Table 2).

**Table 2.**
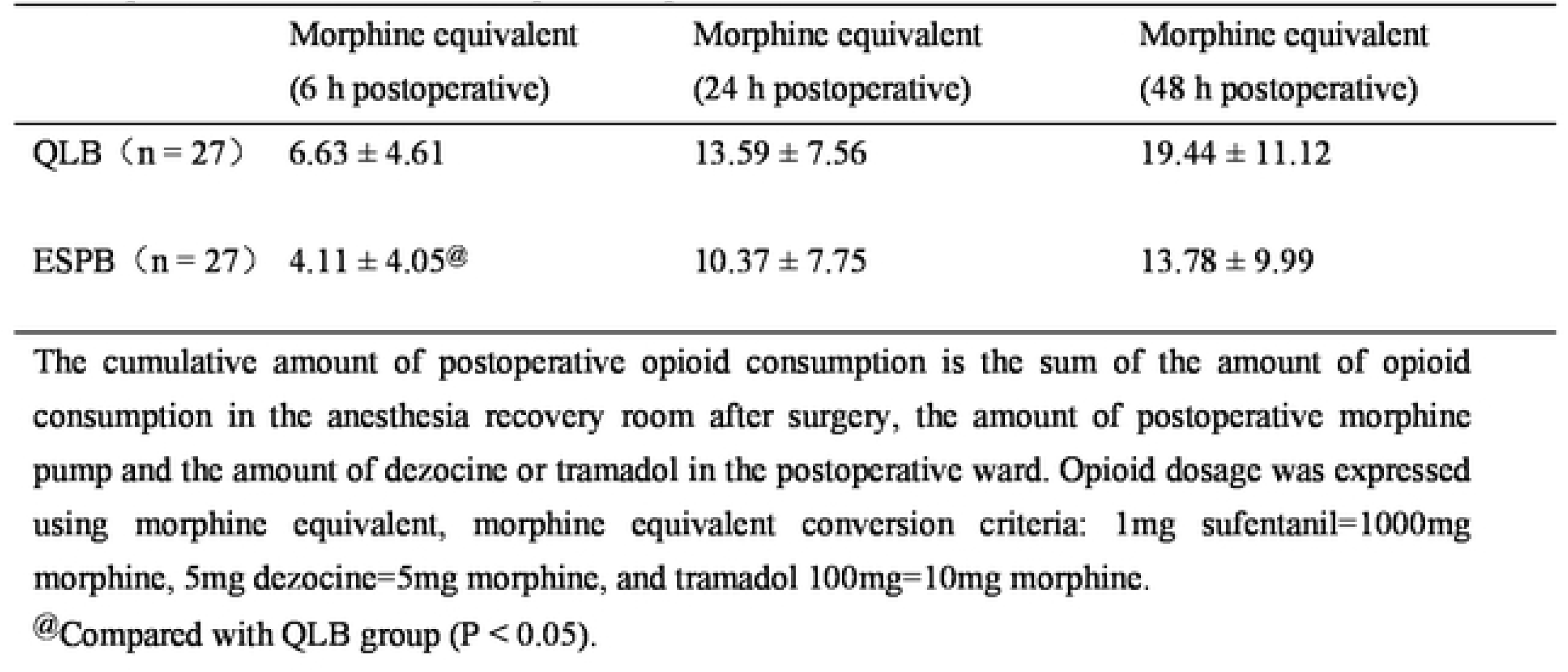
Postoperative cumulative morphine equivalent.

Compared with the QLB group,MAP decreased significantly at T1 in the ESPB group (P <0.05). There was no significant difference in MAP at other time points (P> 0.05).There were no statistically significant differences in HR(P>0.05)(Table 3).

**Table 3.** MAP and HR comparison at each time point.

|  |  | T0 | T1 | T2 | T3 | T4 |
| --- | --- | --- | --- | --- | --- | --- |
| MAP<br>(mmHg) | Q (n=27) | 99.04±11.81 | 85.41±7.47 | 92.22±11.45 | 97.04±10.71 | 98.22±12.01 |
|  | E (n=27) | 100.37±13.76 | 80.63±4.94@ | 89.48±9.61 | 97.48±13.77 | 99.89±10.67 |
| HR<br>(times / min) | Q (n=27) | 78.78±9.46 | 63.00±9.52 | 68.41±13.22 | 84.15±12.36 | 86.44±9.91 |
|  | E (n=27) | 79.78±11.27 | 67.15±10.78 | 70.22±12.89 | 85.11±12.57 | 86.65±16.64 |

Both the QLB and ESPB groups were compared using the Mann-Whitney U test. T0=anesthesia preparation room; T1=5min before surgical skin incision; T2=5min after surgery; T3=5min before tracheal extubation; T4=5min after tracheal extubation; T5=when leaving PACU.
@ Compared with QLB group (P < 0.05).

There was no statistical difference in the use of vasoactive drugs including ephedrine, nooxyadrenaline, esmolol, nicardipine and atropine (P>0.05)(Table 4).

**Table 4.**
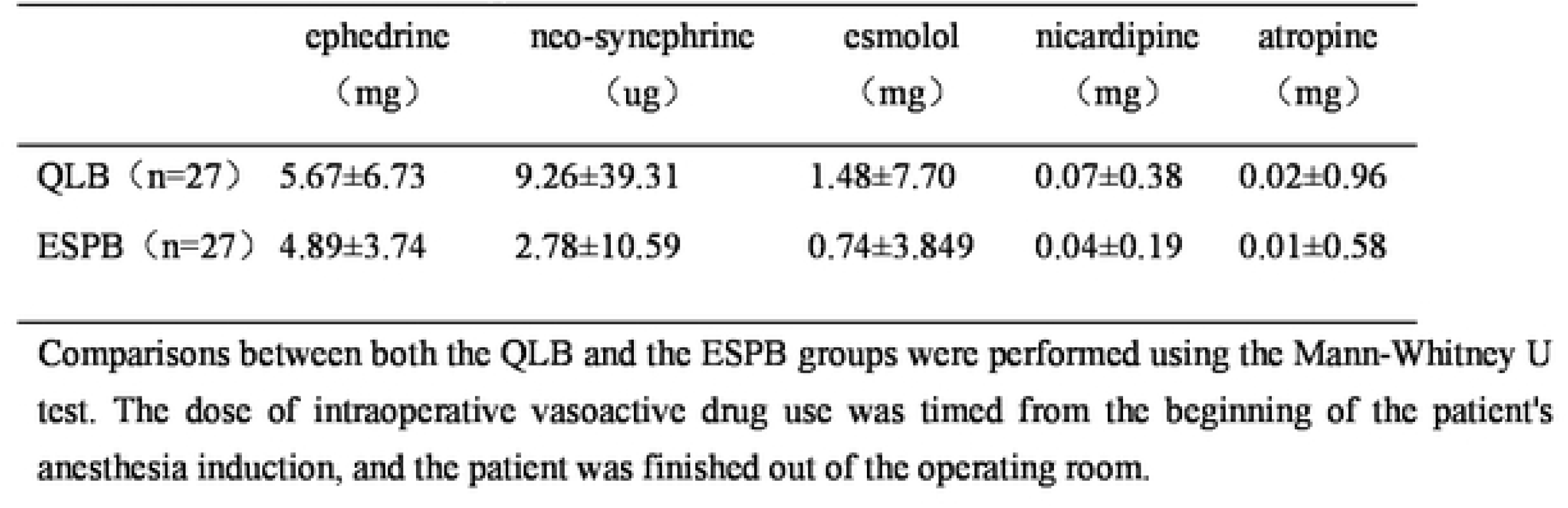
The use of vasoactive drugs.

Compared with the QLB group, resting NRS pain scores were significantly lower at 0.5h (P <0.05). All other time points showed lower resting NRS pain scores, but no statistical difference (P> 0.05). Patients in the EPSB group had lower cough NRS pain score at 0.5h compared with the QLB group, but there was no statistical difference in cough NRS pain scores at all postoperative time points (P> 0.05). In the EPSB group, the number of morphine pump presses decreased at 6h and 24h (P <0.05) and no difference at other time points (P> 0.05) (Table 5).

**Table 5.** Comparison of NRS scores and PICA presses.

|  |  | 0.5h | 6h | 24h | 48h |
| --- | --- | --- | --- | --- | --- |
| Resting NRS pain score | QLB (n=27) | 1.74±1.20 | 1.41±1.22 | 1.30±1.03 | 0.70±0.87 |
|  | ESPB (n=27) | 1.04±0.94@ | 1.07±0.78 | 0.78±0.89 | 0.59±0.69 |
| Cough NRS pain score | QLB (n=27) | 2.30±1.38 | 2.07±1.24 | 2.37±1.04 | 1.52±1.09 |
|  | ESPB (n=27) | 1.74±1.35 | 2.01±0.92 | 1.89±0.89 | 1.56±0.89 |
| Morphine pump presses<br>(times) | QLB (n=27) | 0.85±0.99 | 3.37±2.22 | 7.04±4.47 | 9.37±5.97 |
|  | ESPB (n=27) | 0.52±0.58 | 1.67±1.44@ | 4.52±2.85@ | 6.67±4.32 |

Mann-Whitney U test was used for comparison between QLB and EPSB groups.
@ Compared with the QLB group was statistically significant(P <0.05).

Compared with the QLB group, the operation time of the nerve block was 2.09±0.68min and the depth of the nerve block puncture was 2.14±0.39cm in the ESPB group. Significant comparative difference (P <0.05)(Table 6).

**Table 6.**
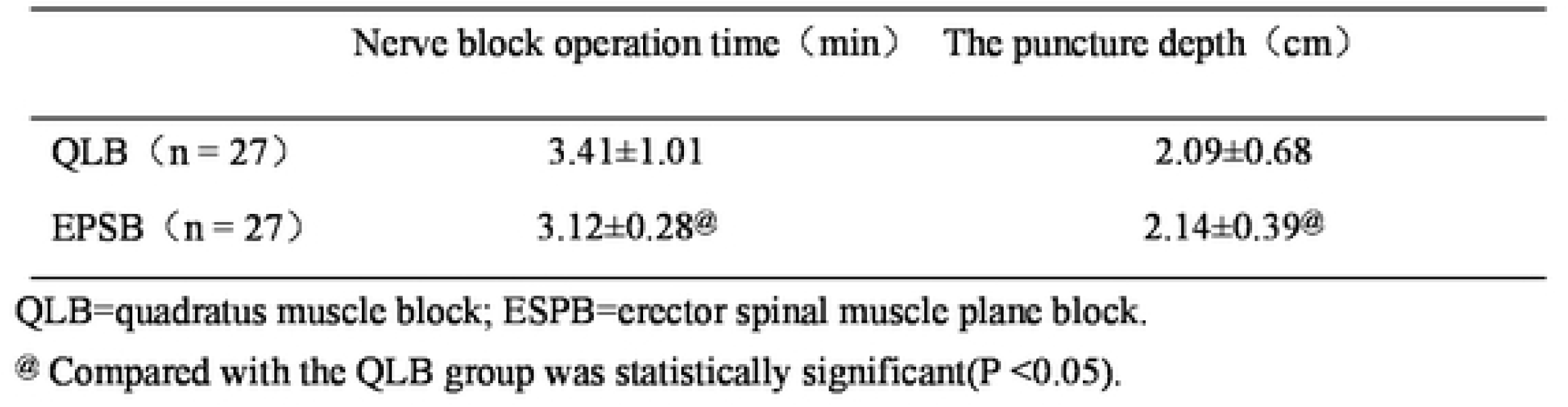
Comparison of operating time and puncture depth.

Compared with QLB group, QoR-15 score decreased at 24 h, and all differences were significant(P<0.05)(Table 7).There were no statistical difference in postoperative nerve block-related complications (puncture site hematoma,puncture site infection, local anesthetic poisoning,respiratory depression,nausea and vomiting,lower limb weakness etc) between the two groups (P> 0.05)(Table 8).On a return visit 1 week after surgery,one patient in the QLB group had mild numbness and discomfort in the inguinal area of the nerve block.

**Table 7.** Quality of anesthesia recovery after surgery.

|  | QLB (n = 27) | ESPB (n = 27) | p value |
| --- | --- | --- | --- |
| QoR-15 (24 h) | 118.85±6.77 | 124.78±3.40 | 0.001@ |
| QoR-15 (48 h) | 125.52±8.20 | 128.04±1.77 | 0.070 |

Both group comparisons were performed using the Mann-Whitney U test.
@ Compared with the QLB group was statistically significant(P <0.05).

**Table 8.**
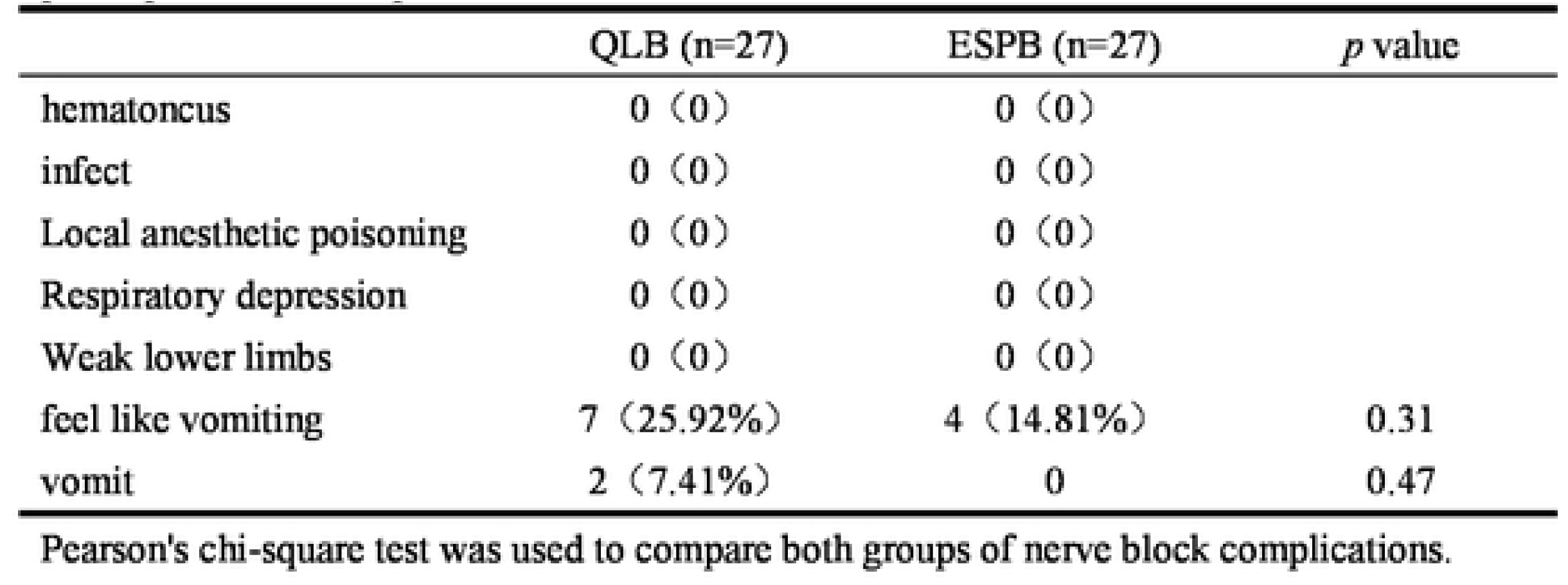
postoperative complications.

No statistical differences were found in the postoperative time out of bed and time to first flatus between the two groups.In EPSB patients, the postoperative hospital stay was shorter than that in QLB patients (P <0.05) (Table 9).

**Table 9.**
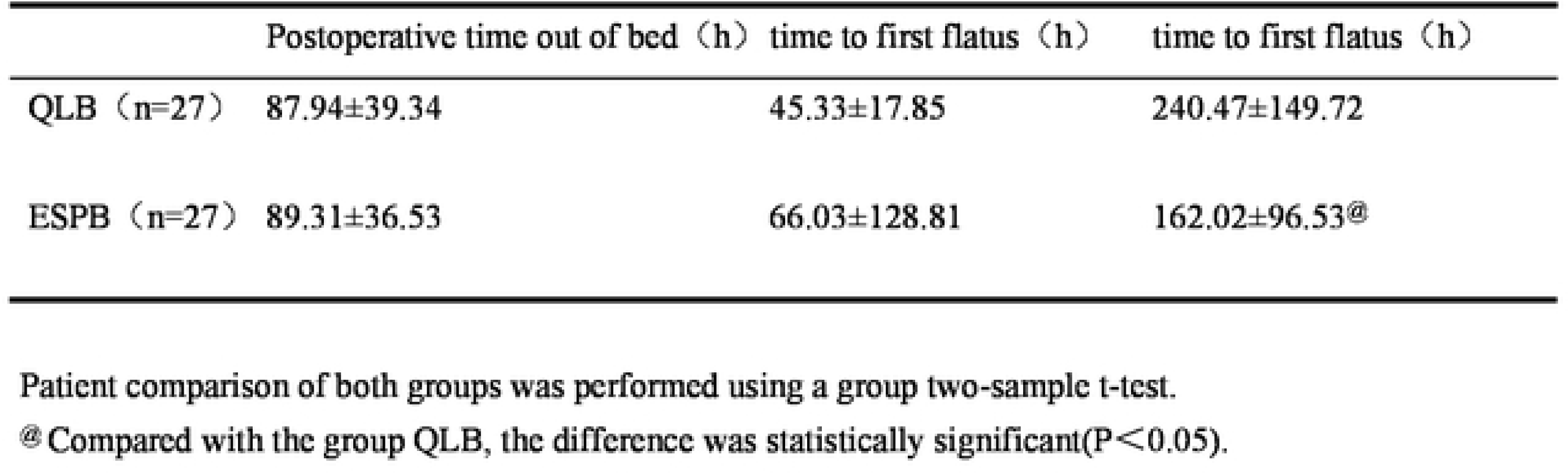
Comparison of postoperative recovery time.

## Disscussion

We conducted a randomized controlled trial in which 54 patients received QLB or ESPB before LN to investigate the use of contrast ultrasound-guided these two nerve block methods in LN.The study found that ultrasound-guided ESPB puncture was shallower and shorter than QLB puncture. At T1 time points, patients with ESPB had a higher incidence of hypotension,but did not significantly increase the intraoperative dose of vasoactive drugs. Patients in the ESPB group had significant improvements in cumulative morphine equivalent consumption at 6h after surgery, resting NRS pain score at 0.5h after surgery, morphine pump compression at 6h and 24h after surgery,24h QoR-15 score after surgery and the length of hospital stay was shortened.

ESPB is a new ultrasound-guided nerve block technique emerging in recent years, which was first proposed by Forero[14] and used in the treatment of neuropathic pain in 2016. A randomized controlled non-inferiority study [15] of ESPB or TPVB block in T9 plane before LN, both using 0.5% ropivacaine 25ml, showed that the postoperative analgesia was not inferior to TPVB after ESPB. Sahin et al[16] treated T10 plane ESPB with bupivacaine 0.25% bupivacaine 30ml before LN.The study results showed a significant improvement in NRS pain score, opioid consumption and quality of recovery score within 20h after surgery in the intervention group. All results of these studies indicate that ESPB has better analgesic effect in LN surgery.

QLB was first proposed by Blanco in 2007[17], and was subsequently further studied and improved.Currently, there are four approach methods for QLB[18] which is widely used in clinical practice.The lateral approach (QLB-I),posterior approach (QLB-II),anterior approach (QLB-III) and muscular approach (QLB-IV) were respectively. QLB-II was used in this study[18], and the drug injection position was located in the area of the lumbar fascia triangle. The local anesthetic could easily spread along the thoracolumbar fascia, and the block segment could reach T7∼L1. Compared with other methods, this method is more superficial, the injection position has clearer ultrasound imaging, and the quadratus muscle between the needle tip and the peritoneum, which is safer to operate. Li et al [19] used QLB-I and QLB-II after LN, both using 0.4% ropivacaine 30ml, the study results showed that the two blocks did not reduce opioid consumption, but improved analgesia within 24 hours after surgery. Zhu et al [20] performed QLB-II before LN and injected 30ml of 0.375% ropivacaine after reaching the block site. The study showed that the cumulative consumption of sufentanil within 12 hours after surgery was lower in the intervention group, and the quality of recovery was higher at 48 hours after surgery.

In this study, ultrasound-guided ESPB was easier to achieve and operate than QLB puncture, which may be related to the anatomical localization of the way we block the needle. The anatomy of quadratus muscle varies greatly by individuals, such as age, obesity, anatomical structural variation, quadratus muscle atrophy in elderly patients, unclear display of deep structure in obese patients and anatomical structural variation will increase the difficulty of quadratus muscle block. However, the erector spinal muscle block was localized to the T10 transverse process, and the drug was injected directly on the surface of the transverse process, with a superficial location and low difficulty in puncture. In addition, we chose the QLB-II approach to use in-plane injection and the ESPB to use out-of-plane injection, which may also be related to our ultrasound-guided nerve block injection method.

The ESPB group had significantly lower MAP at T1 compared with the QLB group. Studies have shown that [21] performed ESPB at the T7 level in 10 volunteers, the drug consisted of 30ml 2.5mg/ml ropivacaine and 0.3ml gadolinium, the results showed that 9/10 volunteers spread to the paravertebral space, 8/10 to the foramina, 4/10 to the epidural, and one volunteer developed extensive epidural diffusion and diffusion of the contralateral epidural and interforamen. Schwartzmann[22] et al in an observational MRI study of local anesthetic (29.7mL 0.25% bupivacaine and 0.3mL gadolinium) in six pain patients, erector, spinal dorsal branch, intercostal space, and foramina, two of which spread to the epidural space. Another case report [23] found that the patient had T8 level ESPB and injected 0.5% ropivacaine 20ml. The patient developed severe hypotension and blocked segment T2-L5.The occurrence of intraoperative hypotension may be related to the extensive epidural spread of local anesthesia.Some studies have shown that the QLB-II block effect is less precise, rarely blocked to the thoracic paravertebral space,affected by large individual differences.In a study[24],three different approaches injected 0.375% ropivacaine in 18ml and 2mL contrast medium,then performed 3D computed tomography (3D-CT) to evaluate the distribution of the injection, which showed that only QLB-III occasionally spread to the thoracic side, whereas QLB-I and QLB-II only spread in the transverse fascia plane and posterior muscle. These findings could explain that the significant reduction of MAP at T1 in the ESPB group compared with the QLB group may be related to the extensive epidural diffusion of local anesthetic drugs.

Compared with the QLB group, resting NRS pain scores at 0.5h after surgery, the number of morphine pump presses at 6h and 24h,cumulative morphine equivalent consumption at 6h were significantly improved in the ESPB group.This is not entirely consistent with a study by Aygun [25] et al, in which QLB-II and ESPB in 40 patients undergoing laparoscopic cholecystectomy (LC) found a significant difference in NRS resting/cough pain scores for the first hour after ESPB,but found no significant difference in opioid consumption,which may be related to lower pain after LC.In another randomized controlled study[26],patients underwent QLB-II and ESPB block after cesarean section,and both received 0.3mL/kg 0.25% bupivacaine.The results showed that QLB-II or ESPB had similar analgesic effect after cesarean section, which may be related to L_3-4_ subarachnoid block and 1.8-2.2ml 0.5% bupivacaine reduced postoperative pain and masked the evaluation of nerve block effect in the two groups.

Compared with the QLB group,the QoR-15 score varied significantly after surgery at 24h in the ESPB group,and this result was also confirmed in the randomized controlled study of Moorthy et al[27].In this study, ESPB and paravertebral block (PVB) were performed after chest surgery and inserted for continuous local anesthetic infusion, QoR-15 score was significantly higher in ESPB patients. Another study showed[28] preoperative QLB in open gastrointestinal surgery, could improve the QoR-15 score at 48 hours and improve the early postoperative analgesia.The higher QoR-15 score after ESPB, we speculate, may be related to the lower postoperative pain score.

There were no statistical differences in the postoperative nerve block-related complications (puncture site hematoma, puncture site infection, local anesthetic poisoning, respiratory depression, nausea and vomiting, lower limb weakness on the affected side) between the QLB and ESPB groups. Some studies reported[29,30]patients developed double lower limb muscle weakness and shoulder radiation pain after undergoing QLB or ESPB, but no such complications were reported in our study. The time of first implantation; there was no statistical difference in first discharge time, and the postoperative hospital stay time of Group ESPB was shorter compared with the QLB group, which is consistent with Yao et al [31].

There are some limitations to our study. First, after nerve block, we used the alcohol contact skin cold sensation regression method to evaluate the block effect. However, due to the limited time of nerve block operation before surgery, some patients only evaluated the block effect, and failed to record the range of the nerve block plane. Second, the surgeon requires patients who undergo laparoscopic total renal resection or radical resection to get out of bed as early as possible after surgery, while the risk of rebleeding in laparoscopic partial renal resection suture is greater, and the first implantation time is relatively conservative. Third, this study was only a single-center and the study sample size was limited. Finally, considering that patients had good postoperative analgesia, no blank control group was established in this study.

## Conclusion

Compared with the operative anterior muscle block, the erector spinal muscle plane block has some advantages in terms of analgesia and recovery quality after laparoscopic nephrectomy, and shows an opioid thrifty effect after surgery. Further attempts can be made to explore the optimal concentration dose of fixed local anesthetic drugs.

## Data availability statement

The datasets used and/or analysed during the current study available from the corresponding author on reasonable request.

## Assistance with the study

we would like to thank Dr.Guangmin Xu, Senior consultant Anaesthesiologist, for his assistance with the study and Dr.Peng Su for statistical assistance.

## Presentation

none.

